# The Ecology of Antihypertensives in the United States, 1997-2017

**DOI:** 10.1101/2020.04.01.20050146

**Authors:** Michael E. Johansen, Joshua D. Niforatos, Jeremey B. Sussman

**Affiliations:** Grant Family Medicine, OhioHealth, Columbus, OH; Heritage College of Osteopathic Medicine at Ohio University, Dublin, OH; Department of Emergency Medicine, The Johns Hopkins University School of Medicine, Baltimore, MD; Department of Internal Medicine, Division of General Medicine, University of Michigan, Ann Arbor, MI, USA

## Abstract

**Background:** Antihypertensives are the most used medication type in the United States, yet there remains uncertainty about the use of different antihypertensives. We sought to characterize use of antihypertensives by and within medication class(es) between 1997-2017.

**Methods:** A repeated cross-sectional study of 493,596 adult individuals using the 1997-2017 Medical Expenditure Panel Survey (MEPS). The Orange Book and published research were used for adjunctive information. The primary outcome was the estimated use by and within anti-hypertensive medication class(es).

**Results:** The proportion of individuals taking any antihypertensive during a year increased from 1997 to the early 2010’s and then remained stable. The proportion of the population taking 2 or more medications declined from 2015-2017. The proportion of adults using angiotensin II receptor-blockers (ARBs) and dihydropyridine calcium channel-blockers (CCBs) increased during the study period, while angiotensin-converting enzyme inhibitors (ACE-Is) increased until 2010 after which rates remained stable. Beta-blocker use was similar to ACE-Is with an earlier decline starting in 2012. Thiazide diuretic use increased from 1997-2007, leveled off until 2014, and declined from 2015-2017. Non-dihydropyridine CCBs use declined throughout the study. ACE-Is, ARBs, CCBs, thiazide diuretics, and loop diuretics all had one dominant in-class medication. There was a clear increase in the use of losartan within ARBs, lisinopril within ACE-Is, and amlodipine within CCBs following generic conversion. Furosemide and hydrochlorothiazide started with and maintained a dominant position in their classes. Metoprolol use increased throughout the study and became the dominant beta-blocker, while atenolol peaked around 2005 and then declined thereafter.

**Conclusions:** Antihypertensive classes appear to have a propensity to equilibrate to an individual medication, despite a lack of outcomes based research to compare medications within a class. Future research could focus on comparative effectiveness for within-class medications early in the life cycle of therapeutics that are probable to have wide spread use.

## Introduction

Over one-fifth of adults in the United States take an anti-hypertensive medication, which makes them the most used type medications in the United States.^1,2^ Antihypertensives are also among the best-studied of all drugs, with at least 464,000 people randomized into antihypertensive clinical trials by 2003.^3^ They are also the topic of multiple high-profile clinical practice guidelines and performance measures.^4,5^ Which anti-hypertensive drugs are used, why they are used, and ensuring that effective clinical research exists to use the most effective drugs possible, could have a large public health impact. But which drugs are used and for what reason is not always known.

In fact, there are relatively few studies of the determinants of the use of specific classes of antihypertensive medications and even fewer examining which drugs are used within specific classes.^1,6–9^ Few trials have directly compared drugs within classes. As a result, guidelines and previous studies have mostly referred to medications as classes rather than unique individual compounds. However, indirect comparisons have provided evidence that there could be important intra-class differences.^10–13^ Therefore, understanding the relationships between evidence, guidelines, and both between-class and within-class clinical use could help guide both implementation and research going forward.

Given this context, the primary goal of this study is to gain a better understanding of the between- and within-class trends in anti-hypertensive medication use over the last 20 years. Due to the complexity of individual decision-making, this descriptive study compiles a hypothesis generated list of observations based on an understanding of the literature. We hypothesized that between-class trends occur in relation to landmark studies, society guidelines, and cost of medications, whereas within-class trends of antihypertensive agents occur in relation to availability, generic conversion, and trial level evidence.

## Methods

### Study Participants

A repeated cross-sectional study of the Medical Expenditure Panel Survey (MEPS) was used to describe trends in antihypertensive medication use over time.^14^ MEPS is sponsored by the Agency of Healthcare Research and Quality (AHRQ) and is representative of the non-institutionalized population of the United States. Each year of the survey is comprised of two overlapping cohorts, which are interviewed five times over the 2 years. The survey includes information regarding demographic, socioeconomic, and prescription medication utilization. All adult individuals included in the survey between 1997-2017 were included in the analysis.

### Study Variables

Anti-hypertensive medications were identified through a combination of therapeutic classification and prescription medication name. The survey uses numerous methods to achieve accurate reporting of prescription medications including contacting pharmacies.^15–17^ Previous studies have validated the accuracy of chronic medication reporting in MEPS.^18^ Included classes in the study were angiotensin converting enzyme inhibitor (ACE-I), thiazide diuretic, angiotensin receptor blocker (ARB), beta-blocker, loop diuretic, dihydropyridine calcium channel blockers (CCB), non-dihydropyridine CCB, aldosterone receptor antagonists, hydralazine, and clonidine. Other drug classes were not studied given low levels of use and/or numerous therapeutic uses across numerous categories. Within these classes, individual medications were identified by name. Combination medications were reported in applicable categories. An individual was identified as a medication user if he or she had any use of the medication during a given year. Generic availability of medications was identified as the first approved Abbreviated New Drug Application (ANDA) through the Food and Drug Administration (FDA).

### Analysis

Complex survey weights were used in all analyses to makes these analyses representative of the non-institutionalized population of the United States. 95% confidence intervals are reported in the figures.

To contextualize identified trends, we explored landmark randomized clinical trials, germane societal guidelines, and generic conversions of name-brand medications. We opted to include a selection of key clinical practice guidelines issued by the (or by panel members of) Joint National Committee on Prevention, Detection, Evaluation, and Treatment of High Blood Pressure.^19^ Randomized clinical trials were identified from a recent history of hypertension.^3,20^

The OhioHealth Institutional Review Board ruled the study exempt. All analyses used STATA 15 (College Station, Tx).

## Results

Between 1997-2017, 493,596 individuals were identified. We identified 43 relevant anti-hypertensive medications among 10 classes (Table 1). All but 4 of these drugs were approved before our study dataset began in 1997, but 26 achieved generic status between 1997 and 2016. The newest drugs were 3 angiotensin receptor blockers and nebivolol.

**Table 1.**
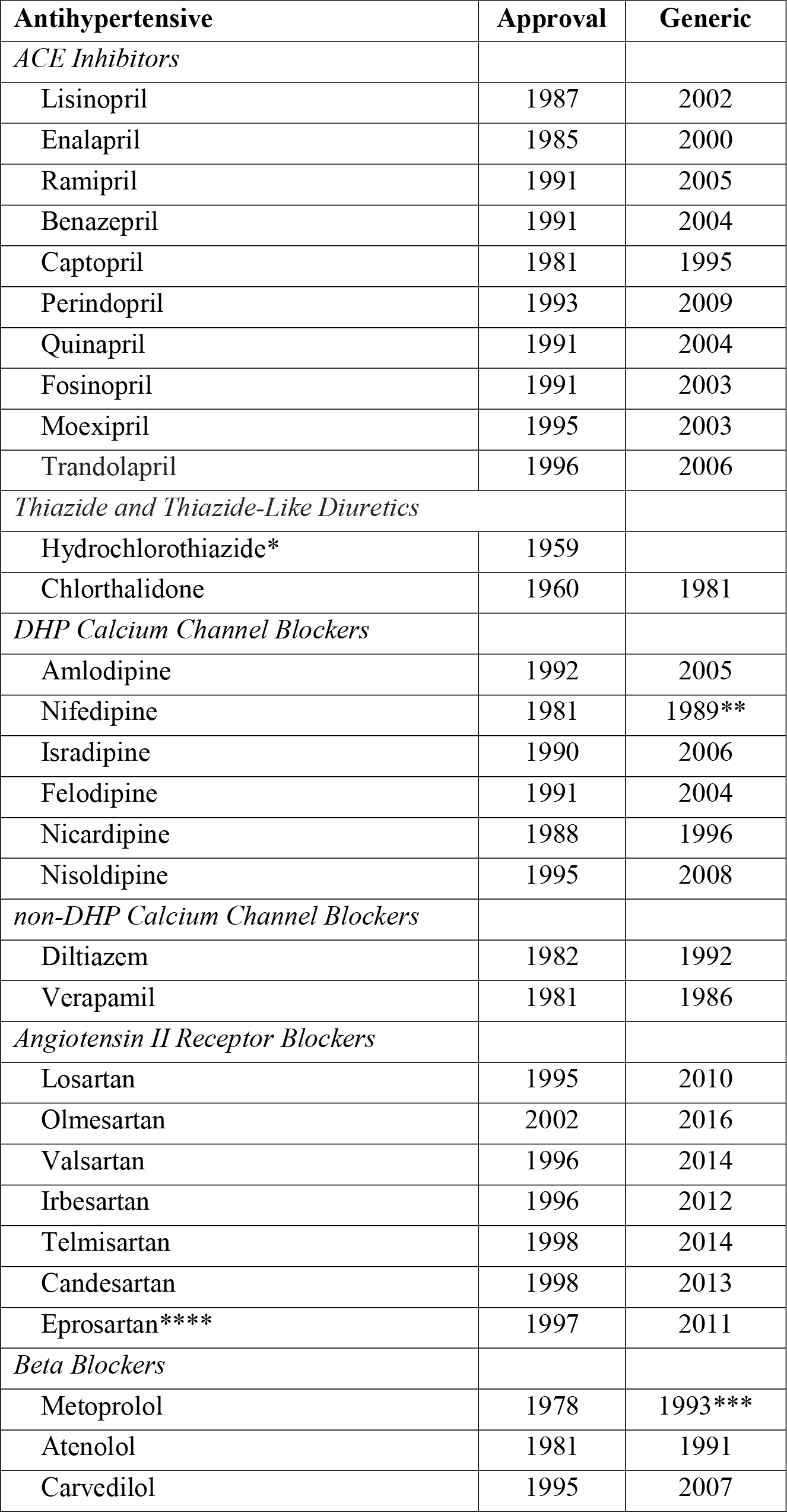
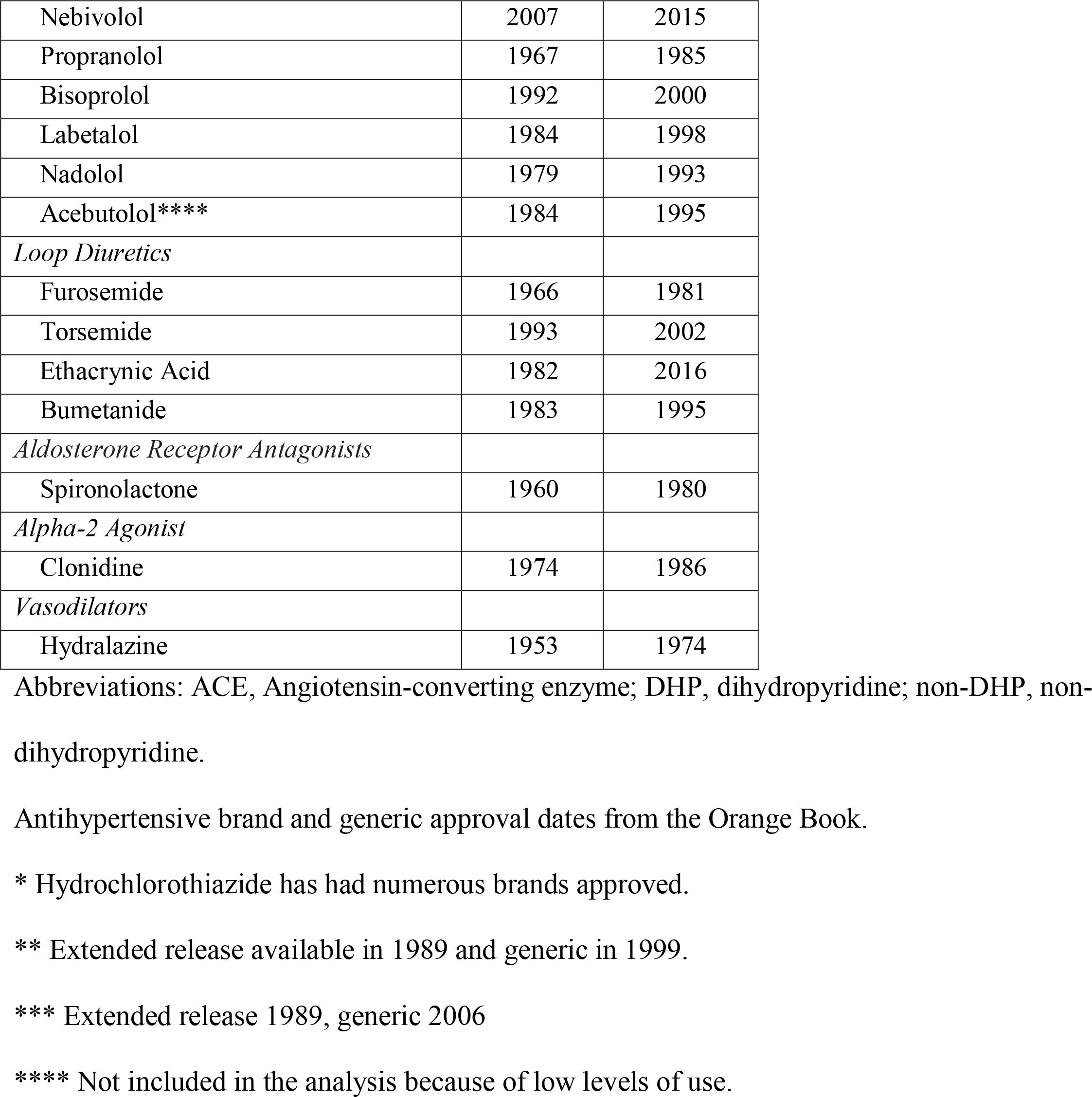
**Antihypertensive Medications Brand and Generic Approval Dates**

The proportion of individuals taking antihypertensive medication during a year increased from 1997 to the early 2010’s (Figure 1). The proportion taking 2, 3, and 4 medications all followed a similar pattern. However, from 2015-2017, the proportion of the population taking 2 or more medications declined, while the number taking one increased.

**Figure 1.**
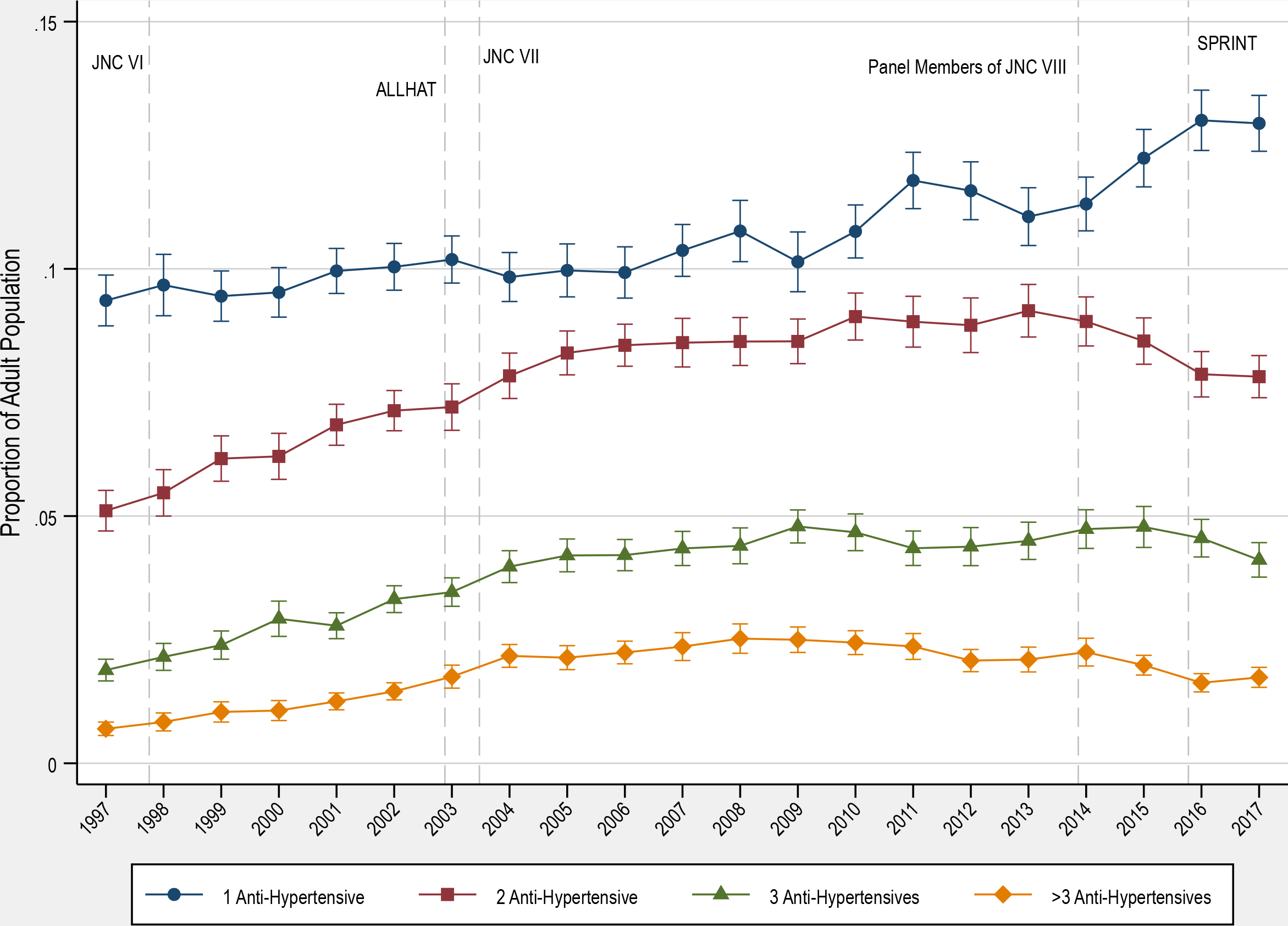
Proportion of the Adult Population on Antihypertensives. Figure 1 illustrates the proportion of the adult population who are on a particular number of anti-hypertensive medication classes between 1997-2017. A user was defined by the report of any medication in a medication class. Dashed vertical lines represent publication of landmark clinical trials or publication date of clinical practice guidelines. Data from the 1997-2017 Medical Expenditure Panel Survey were used. Brackets represent 95% confidence intervals.

Use of drugs within classes changed substantially over time (Figure 2). Use of ARBs and dihydropyridine CCBs increased substantially during the study period. The proportion of the population who used ACE-Is increased until 2010 after which rates remained stable before a small decline in 2016-2017. Beta-blocker use was similar to ACEIs with an earlier decline starting in 2012. Thiazide diuretic use increased from 1997-2007, leveled off until 2014, and declined from 2015-2017. Non-dihydropyridine CCBs use declined throughout the study period. Loop diuretic, spironolactone, and clonidine rates of use were largely unchanged, while the proportion of people using hydralazine increased slightly. The relative relationship between medication classes did not change dramatically after the publication of the ALLHAT trial. Within classes there was a striking tendency towards one drug developing one dominance in the class, but when the drug became dominant varied greatly (Figure 3). The thiazide diuretic hydrochlorothiazide and the loop diuretic furosemide were dominant before 1997 began and became more so by 2017. The ACE-I lisinopril, ARB losartan, beta-blocker metoprolol, and CCB amlodipine all became dominant. Within-class dominance sometimes followed generic conversion, but sometimes predated it. The dominant drug was the first introduced in 3 of 6 major classes.

**Figure 2.**
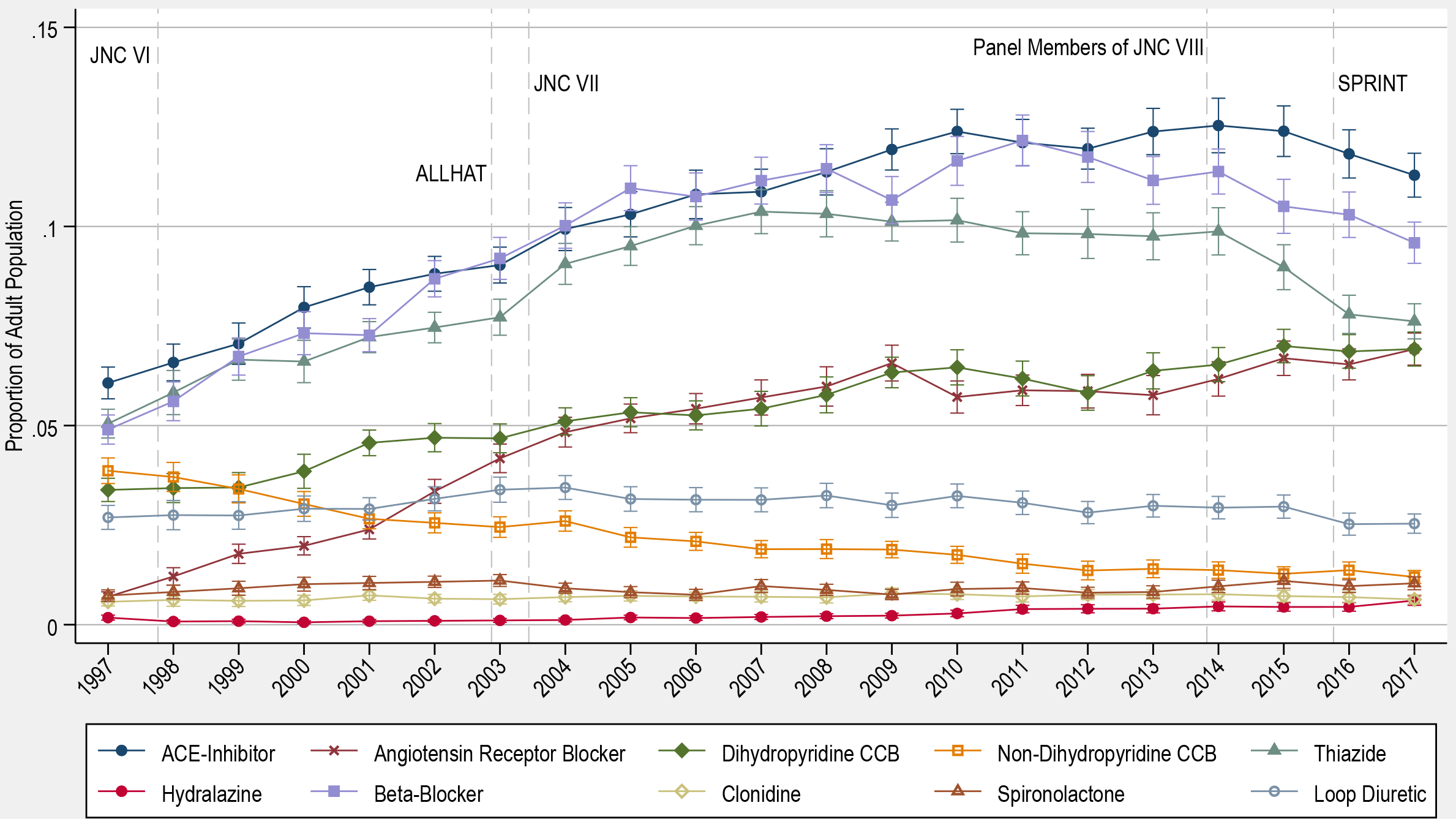
Proportion of Adult Population using Anti-Hypertensive from Medication Classes. Abbreviations: ACE, Angiotensin-converting enzyme; CCB, calcium channel blocker; JNC VI, Sixth Report of the Joint National Committee; JNC VII, Seventh Report of the Joint National Committee; JNC VIII, Eight Joint National Committee; ALLHAT, Antihypertensive and Lipid-Lowering Treatment to Prevent Heart Attack Trial; SPRINT, Systolic Blood Pressure Intervention Trial. Figure 2 depicts the proportion of the adult population who reported use of an anti-hypertensive from different medication classes during a year using the 1997-2017 Medical Expenditure Panel Survey. Dashed vertical lines mark important clinical trials or clinical practices guidelines that could have influenced use of different medications. Brackets represent 95% confidence intervals.

**Figure 3.**
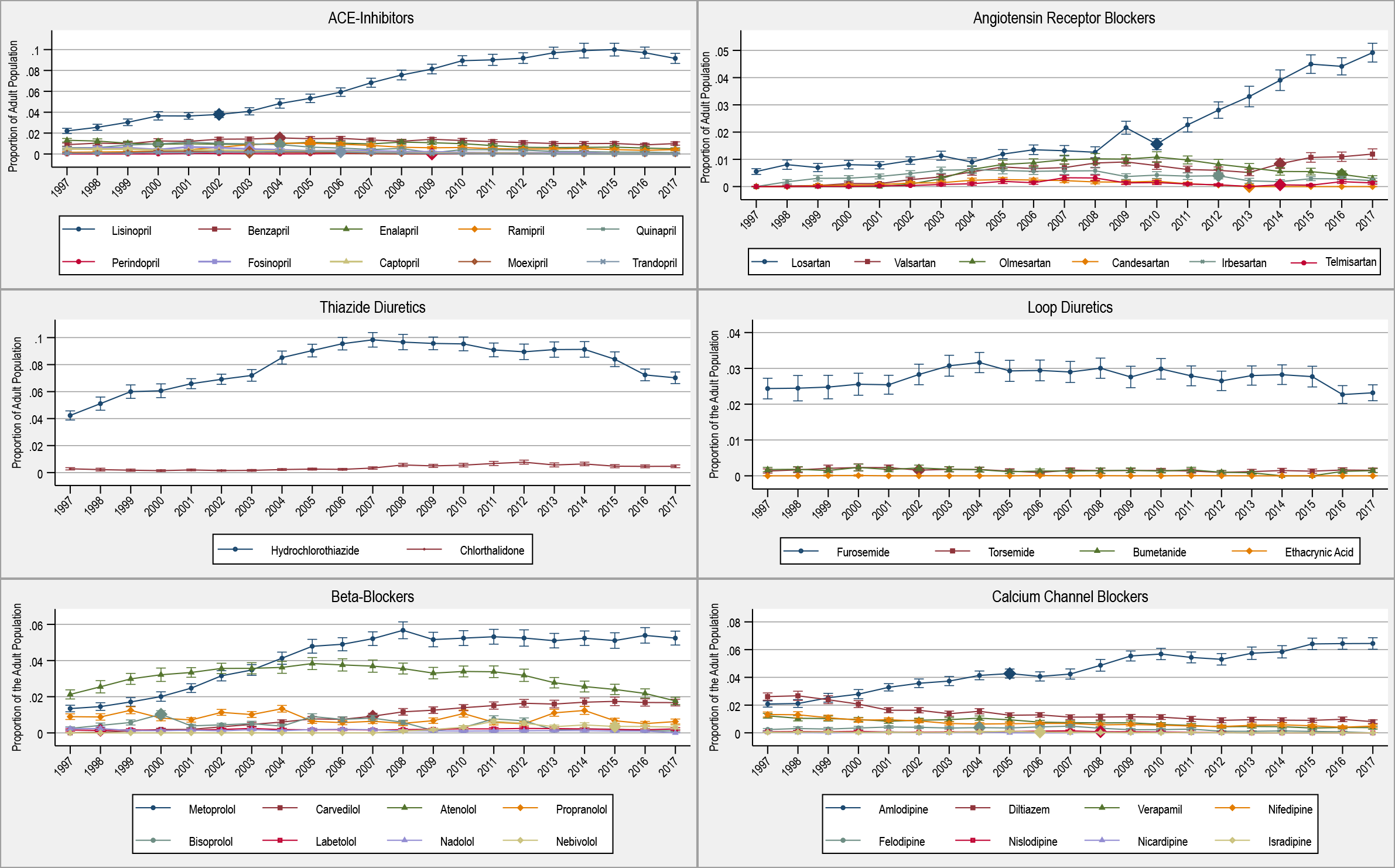
Proportion of Adult Population using Different Medications by Medication Class. Abbreviations: ACE, Angiotensin-converting enzyme. Figure 3 provides information on the proportion of the adult population who reported use of individual medication organized by medication class by year. The 1997-2017 Medical Expenditure Panel Survey was used. The Y-axes represents the proportion of the adult population. Large diamonds represent the year a medication had an approved generic (also see Table 1). Brackets represent 95% confidence intervals.

## Discussion

Our findings show a mismatch between within-class and between-class use choices of anti-hypertensive medications. Within classes, we found that most antihypertensive medication classes eventually equilibrate to one dominant medication. There was no clear evidence that the medication that became dominant did so for reasons of formal studies demonstrating superiority within the class. The dominant medication was not consistently associated with early entry or early generic availability. Drugs with some evidence of comparative advantage (i.e. chlorthalidone, torsemide)^21–25^ did not gain dramatically change in use, while atenolol did appear to decrease in use with less robust evidence.^26-28^

However, between-class medication use was partly associated with key studies like SPRINT, and clinical practice guidelines.^29^ As guidelines until 2014 recommended generally intensifying treatment, overall use followed. With the decline in treatment intensity recommended in the 2014 clinical practice guidelines,^19^ there was a small change in clinical practice. Treatment did not change dramatically after either ALLHAT or the SPRINT trials. Medications with more negative harm profiles had declining use over time.^30-32^ Beta-blockers and CCBs had more movement over the course of the study, which might have been related to mechanistic differences and alternative uses of individual medications.

These data do raise the concern that if small-to-moderate differences in effect or adverse events within classes exist (i.e. phenformin in biguanides or troglitazone in thiazolidinediones), current practice may not recognize those differences. For instance, if one of the numerous ACE-Is lowered overall mortality or disease-oriented outcomes by 10% more than other drugs of that class, it could improve the health or adherence to medications of a population, though would require very large clinical trials to be effective, likely four times the size of the SPRINT trial, assuming otherwise similar populations. Determining when medications and medication classes will justify these studies would be challenging and require foresight. Although a few comparative trials are underway (e.g., ClinicalTrials.gov NCT03978884, NCT03928145, NCT02185417), these obstacles are formidable. Alternatives to classical clinical trials offer some hope, but challenges remain with implementing these types of trials.^33^

A few limitations of this study are worth nothing. First, the high-level (ecological) observational nature of our data makes strong explanatory conclusions inappropriate. We cannot rule out residual confounding, demographic or health changes, costs, or other, unmeasured causes of change in use, including advertising and promotion.^34-37^ Additional limitations include possible under-reporting of medications, a lack of biometric measurements, and potential utilization of certain medications for non-blood pressure related purposes.

## Conclusion

We found that over the last 20 years anti-hypertensive drug classes have tended to equilibrate on one in-class medication. Additionally, there have been shifts in the types and amount of medications used in the population. Future research on comparative effectiveness for within-class medications early in the life cycle that are probable to have wide-spread use might improve health. In the absence of within-class randomized clinical trials, future funding for building the infrastructure for randomized evaluations in the ambulatory setting might be an avenue to do these trials, though these study designs are not without limitations.^30^ Without this renewed focus on within-class drug efficacy, prescribing patterns for antihypertensive medications within-class and between-class may continue to be done in the presence of considerable uncertainty.

## Data Availability

This data is publicly available data.

## References

1. Shah SJ, Stafford RS. Current Trends of Hypertension Treatment in the United States. Am J Hypertens. 2017;30(10):1008–1014. doi:10.1093/ajh/hpx085

2. Fuentes AV, Pineda MD, Venkata KCN. Comprehension of Top 200 Prescribed Drugs in the US as a Resource for Pharmacy Teaching, Training and Practice. Pharm Basel Switz. 2018;6(2). doi:10.3390/pharmacy6020043

3. Law MR, Wald NJ, Rudnicka AR. Quantifying effect of statins on low density lipoprotein cholesterol, ischaemic heart disease, and stroke: systematic review and meta-analysis. BMJ. 2003;326(7404):1423. doi:10.1136/bmj.326.7404.1423

4. Whelton PK, Carey RM, Aronow WS, et al. 2017 ACC/AHA/AAPA/ABC/ACPM/AGS/APhA/ASH/ASPC/NMA/PCNA Guideline for the Prevention, Detection, Evaluation, and Management of High Blood Pressure in Adults: A Report of the American College of Cardiology/American Heart Association Task Force on Clinical Practice Guidelines. J Am Coll Cardiol. 2018;71(19):e127–e248. doi:10.1016/j.jacc.2017.11.006

5. Casey DE, Thomas RJ, Bhalla V, et al. 2019 AHA/ACC Clinical Performance and Quality Measures for Adults With High Blood Pressure: A Report of the American College of Cardiology/American Heart Association Task Force on Performance Measures. J Am Coll Cardiol. 2019;74(21):2661–2706. doi:10.1016/j.jacc.2019.10.001

6. Gu Q, Burt VL, Dillon CF, Yoon S. Trends in antihypertensive medication use and blood pressure control among United States adults with hypertension: the National Health And Nutrition Examination Survey, 2001 to 2010. Circulation. 2012;126(17):2105–2114. doi:10.1161/CIRCULATIONAHA.112.096156

7. Jarari N, Rao N, Peela JR, et al. A review on prescribing patterns of antihypertensive drugs. Clin Hypertens. 2015;22:7. doi:10.1186/s40885-016-0042-0

8. Fang J, Gillespie C, Ayala C, Loustalot F. Prevalence of Self-Reported Hypertension and Antihypertensive Medication Use Among Adults Aged ≥18 Years - United States, 2011- 2015. MMWR Morb Mortal Wkly Rep. 2018;67(7):219–224. doi:10.15585/mmwr.mm6707a4

9. Johansen ME, Yun J, Griggs JM, Jackson EA, Richardson CR. Anti-Hypertensive Medication Combinations in the United States. J Am Board Fam Med JABFM. 2020;33(1):143–146. doi:10.3122/jabfm.2020.01.190134

10. Fretheim A, Odgaard-Jensen J, Brørs O, et al. Comparative effectiveness of antihypertensive medication for primary prevention of cardiovascular disease: systematic review and multiple treatments meta-analysis. BMC Med. 2012;10:33. doi:10.1186/1741-7015-10-33

11. Remonti LR, Dias S, Leitão CB, et al. Classes of antihypertensive agents and mortality in hypertensive patients with type 2 diabetes-Network meta-analysis of randomized trials. J Diabetes Complications. 2016;30(6):1192–1200. doi:10.1016/j.jdiacomp.2016.04.020

12. Qi H, Liu Z, Cao H, et al. Comparative Efficacy of Antihypertensive Agents in Salt-Sensitive Hypertensive Patients: A Network Meta-Analysis. Am J Hypertens. 2018;31(7):835–846. doi:10.1093/ajh/hpy027

13. Wright JM, Musini VM, Gill R. First-line drugs for hypertension. Cochrane Database Syst Rev. 2018;4:CD001841. doi:10.1002/14651858.CD001841.pub3

14. MEPS HC-201 2017 full year consolidated data file. August 2019. https://meps.ahrq.gov/data_stats/download_data/pufs/h201/h201doc.pdf. Accessed February 7, 2020.

15. Cohen J. Design and Methods of the Medical Expenditure Panel Survey Household Component. MEPS Methodology Report No 1.; 1997. https://meps.ahrq.gov/data_files/publications/mr1/mr1.shtml. Accessed February 7, 2020.

16. Cohen JW, Monheit AC, Beauregard KM, et al. The Medical Expenditure Panel Survey: a national health information resource. Inq J Med Care Organ Provis Financ. 1996;33(4):373–389.

17. Cohen JW, Cohen SB, Banthin JS. The medical expenditure panel survey: a national information resource to support healthcare cost research and inform policy and practice. Med Care. 2009;47(7 Suppl 1):S44–50. doi:10.1097/MLR.0b013e3181a23e3a

18. Hill SC, Zuvekas SH, Zodet MW. Implications of the accuracy of MEPS prescription drug data for health services research. Inq J Med Care Organ Provis Financ. 2011;48(3):242–259. doi:10.5034/inquiryjrnl_48.03.04

19. James PA, Oparil S, Carter BL, et al. 2014 evidence-based guideline for the management of high blood pressure in adults: report from the panel members appointed to the Eighth Joint National Committee (JNC 8). JAMA. 2014;311(5):507–520. doi:10.1001/jama.2013.284427

20. Saklayen MG, Deshpande NV. Timeline of History of Hypertension Treatment. Front Cardiovasc Med. 2016;3:3. doi:10.3389/fcvm.2016.00003

21. Carter BL, Ernst ME, Cohen JD. Hydrochlorothiazide versus chlorthalidone: evidence supporting their interchangeability. Hypertens Dallas Tex 1979. 2004;43(1):4–9. doi:10.1161/01.HYP.0000103632.19915.0E

22. Olde Engberink RHG, Frenkel WJ, van den Bogaard B, Brewster LM, Vogt L, van den Born B-JH. Effects of thiazide-type and thiazide-like diuretics on cardiovascular events and mortality: systematic review and meta-analysis. Hypertens Dallas Tex 1979. 2015;65(5):1033–1040. doi:10.1161/HYPERTENSIONAHA.114.05122

23. Roush GC, Ernst ME, Kostis JB, Tandon S, Sica DA. Head-to-head comparisons of hydrochlorothiazide with indapamide and chlorthalidone: antihypertensive and metabolic effects. Hypertens Dallas Tex 1979. 2015;65(5):1041–1046. doi:10.1161/HYPERTENSIONAHA.114.05021

24. Lederle FA, Cushman WC, Ferguson RE, Brophy MT, Fiore Md LD. Chlorthalidone Versus Hydrochlorothiazide: A New Kind of Veterans Affairs Cooperative Study. Ann Intern Med. 2016;165(9):663–664. doi:10.7326/M16-1208

25. Abraham B, Megaly M, Sous M, et al. Meta-Analysis Comparing Torsemide Versus Furosemide in Patients With Heart Failure. Am J Cardiol. 2020;125(1):92–99. doi:10.1016/j.amjcard.2019.09.039

26. Ram CVS. Beta-blockers in hypertension. Am J Cardiol. 2010;106(12):1819–1825. doi:10.1016/j.amjcard.2010.08.023

27. Parker ED, Margolis KL, Trower NK, et al. Comparative effectiveness of 2 β-blockers in hypertensive patients. Arch Intern Med. 2012;172(18):1406–1412. doi:10.1001/archinternmed.2012.4276

28. Carlberg B, Samuelsson O, Lindholm LH. Atenolol in hypertension: is it a wise choice? Lancet Lond Engl. 2004;364(9446):1684–1689. doi:10.1016/S0140-6736(04)17355-8

29. SPRINT Research Group, Wright JT, Williamson JD, et al. A Randomized Trial of Intensive versus Standard Blood-Pressure Control. N Engl J Med. 2015;373(22):2103–2116. doi:10.1056/NEJMoa1511939

30. Furberg CD, Psaty BM, Meyer JV. Nifedipine. Dose-related increase in mortality in patients with coronary heart disease. Circulation. 1995;92(5):1326–1331. doi:10.1161/01.cir.92.5.1326

31. Tedla YG, Bautista LE. Drug Side Effect Symptoms and Adherence to Antihypertensive Medication. Am J Hypertens. 2016;29(6):772–779. doi:10.1093/ajh/hpv185

32. van der Laan DM, Elders PJM, Boons CCLM, Beckeringh JJ, Nijpels G, Hugtenburg JG. Factors associated with antihypertensive medication non-adherence: a systematic review. J Hum Hypertens. 2017;31(11):687–694. doi:10.1038/jhh.2017.48

33. Dal-Ré R, Janiaud P, Ioannidis JPA. Real-world evidence: How pragmatic are randomized controlled trials labeled as pragmatic? BMC Med. 2018;16(1):49. doi:10.1186/s12916-018-1038-2

34. Wilkes MS, Bell RA, Kravitz RL. Direct-to-consumer prescription drug advertising: trends, impact, and implications. Health Aff Proj Hope. 2000;19(2):110–128. doi:10.1377/hlthaff.19.2.110

35. Carey C, Lieber E, Miller S. Drug Firms’ Payments and Physicians’ Prescribing Behavior in Medicare Part D. Natl Bur Econ Res Work Pap. February 2020:1–52.

36. Fleischman W, Ross JS, Melnick ER, Newman DH, Venkatesh AK. Financial Ties Between Emergency Physicians and Industry: Insights From Open Payments Data. Ann Emerg Med. 2016;68(2):153-158.e4. doi:10.1016/j.annemergmed.2016.01.014

37. Vijay A, Gupta R, Liu P, Dhruva SS, Shah ND, Ross JS. Medicare Formulary Coverage of Brand-Name Drugs and Therapeutically Interchangeable Generics. J Gen Intern Med. October 2019. doi:10.1007/s11606-019-05432-6

